# A Postpartum Breast Cancer Diagnosis Reduces Survival in Germline *BRCA* pathogenic variant Carriers

**DOI:** 10.1101/2023.12.21.23300040

**Authors:** Zhenzhen Zhang, Shangyuan Ye, Sarah M. Bernhardt, Heidi D. Nelson, Ellen M Velie, Virginia F Borges, Emma R Woodward, D. Gareth R Evans, Pepper Schedin

**Affiliations:** Division of Oncological Sciences, Oregon Health & Science University, Portland, OR 97239, USA; Knight Cancer Institute, Oregon Health & Science University, Portland, OR 97239, USA; Biostatistics Sharing Resources, Knight Cancer Institute, Oregon Health & Science University, Portland, OR 97239, USA; Department of Cell, Developmental & Cancer Biology, OHSU Knight Cancer Institute, Oregon Health & Science University, Portland, OR 97239, USA; Kaiser Permanente Bernard D. Tyson School of Medicine, Pasadena, CA 91101, USA; Zilber College of Public Health, University of Wisconsin-Milwaukee, Milwaukee, WI 53205, USA; Departments of Medicine and Pathology, Medical College of Wisconsin, Milwaukee, WI 53226, USA; Department of Medicine, Division of Medical Oncology, Young Women’s Breast Cancer Translational Program, University of Colorado Anschutz Medical Campus, Aurora, CO 80045, USA; Manchester Centre for Genomic Medicine, Manchester Academic Health Sciences Centre (MAHSC), Institute of Human Development, St Mary’s Hospital, University of Manchester, Manchester M13 9WL, UK; Prevent Breast Cancer Centre, University Hospital of South Manchester NHS Trust, Wythenshawe, Manchester M23 9LT, UK; Manchester Centre for Genomic Medicine, St Mary’s Hospital, Central Manchester University Hospitals NHS Foundation Trust, Manchester M13 9WL, UK; Manchester Breast Centre, The University of Manchester, Manchester M20 4BX, UK

**Keywords:** postpartum breast cancer, mortality, *BRCA*

## Abstract

**IMPORTANCE:** In young-onset breast cancer, a diagnosis within 5-10 years of childbirth associates with increased mortality. Women with germline *BRCA1/2* pathogenic variants (PVs) are more likely to be diagnosed with breast cancer at younger ages, but the impact of childbirth on mortality is unknown.

**OBJECTIVE:** Determine whether time between recent childbirth and breast cancer diagnosis impacts mortality among young-onset breast cancer patients with germline *BRCA1/2* PVs.

**DESIGN, SETTING, AND PARTICIPANTS:** This prospective cohort study includes 903 women with germline *BRCA1/2* PVs diagnosed with stage I-III breast cancer at ≤45 years of age, between 1950-2021 in the UK.

**MAIN OUTCOMES AND MEASURES:** The primary outcome is all-cause mortality, censored at 20 years post-diagnosis. The primary exposure is time between most recent childbirth and breast cancer diagnosis, with recent childbirth defined as >0-<10 years post childbirth (n=419)], further delineated to >0-<5 years (n=228) and 5-<10 years (n=191). Mortality of nulliparous cases (n=224) was compared to the recent postpartum groups and the ≥10 years postpartum (n=260) group. Cox proportional hazards regression analyses were adjusted for patient age, tumor stage, further stratified by tumor estrogen receptor (ER) and *BRCA* gene status.

**RESULTS:** For all *BRCA* PV carriers, increased all-cause mortality was observed in women diagnosed >0-<10 years postpartum, compared to nulliparous and ≥10 years groups, demonstrating the transient duration of postpartum risk. Risk of mortality was greater for ER-positive cases in the >0-<5 group [HR=2.35 (95% CI, 1.02-5.42)] and ER-negative cases in the 5-<10 group [HR=3.12 (95% CI, 1.22-7.97)] compared to the nulliparous group. Delineated by *BRCA1* or *BRCA2*, mortality in the 5-<10 group was significantly increased, but only for *BRCA1* carriers [HR=2.03 (95% CI, 1.15-3.58)].

**CONCLUSIONS AND RELEVANCE:** Young-onset breast cancer with germline *BRCA* PVs confers increased risk for all-cause mortality if diagnosed within 10 years of childbirth, with risk highest for ER+ cases at >0-<5 years postpartum, and for ER-cases at 5-<10 years postpartum. *BRCA1* carriers are at highest risk for poor prognosis when diagnosed at 5-10 years postpartum. No such associations were observed for *BRCA2* carriers. These results should inform genetic counseling, prevention, and treatment strategies for *BRCA* PV carriers.

**Key Points:** *Question:* Is a postpartum diagnosis an independent risk factor for mortality among young-onset breast cancer patients with germline *BRCA1/2* PVs?

*Findings:* A diagnosis <10 years postpartum associates with higher risk of mortality compared to nulliparous and ≥10 years postpartum cases. Peak risk after childbirth varies for ER-positive (>0-<5 years) vs. ER-negative cases (5-<10 years). *BRCA1* carriers had peak risk of mortality 5-10 years postpartum, with no associations observed for *BRCA2* carriers.

*Meaning:* A breast cancer diagnosis within 10 years of childbirth independently associates with increased risk for mortality in patients with germline *BRCA1/2* PVs, especially for carriers of *BRCA1* PVs.

## Introduction

In the United Kingdom^1^ and the United States^2^, breast cancer diagnosed at age 45 years and younger (young-onset) accounts for approximately 10% of all newly diagnosed invasive breast cancer cases. The incidence of young-onset breast cancer is even higher in other countries, accounting for approximately 19% of all newly diagnosed invasive breast cancer cases worldwide^1^. Further, the incidence trend of young-onset breast cancer has been gradually increasing worldwide for decades^3–6^. This rising incidence is likely unrelated to increased mammographic detection, as the vast majority of cases are too young for routine screening^7,8^.

Rather increased incidence appears due, at least in part, to changes in reproductive factors, including pregnancies occurring at older ages^9^. Although overall treatment has improved outcomes for breast cancer patients at all ages^10^, those with young-onset breast cancer continue to experience elevated mortality rates and have had only modest improvements in treatment efficacy^11^. Importantly, compared to later-age onset breast cancer, young-onset breast cancer is enriched with poor prognostic tumor features^12–16^ and associates with higher mortality^6,11,15,17,18^

An emerging body of work finds the postpartum period as a high-risk window for initiation of new cancers and/or the rapid progression of sub-clinical lesions to cancers with metastatic phenotypes^19–23^. Meta-analyses of young-onset breast cancer showed a postpartum diagnosis up to 10 years following childbirth consistently associates with increased risk of distant metastasis and death^19,20,24^. These breast cancers are defined as postpartum breast cancer (PPBC)^25^. Given that proximity to recent childbirth being is such a strong predictor of breast cancer metastasis and survival in the general population ^19,20,22–24,26–30^, the question of whether women with hereditary pathogenic variants (PVs) in breast cancer pre-disposing genes have similarly poorer prognosis merits investigation. Of the approximately 2.3 million women worldwide diagnosed with breast cancer each year^31^, 5%-6% are due to hereditary gene PVs, with *BRCA* PVs being dominant, accounting for approximately half of inherited cases ^32,33^. PVs in *BRCA1* and *BRCA2* genes were discovered in 1994^34^ and1995 ^35^, respectively, and both genes encode tumor suppression proteins directly linked to homologous recombination repair of DNA^36^. The risk of developing breast cancer is ∼72% for *BRCA1* and ∼69% for *BRCA2* PV carriers^37^, while in the general population, the lifetime risk of developing breast cancer is 13%. The peak incidence for *BRCA* carriers is also younger than the general population, occurring in the 41 to 50-year age group for *BRCA1* carriers and 51- to 60-year age group for *BRCA2* carriers^37^.

To better understand the impact of recent childbirth on prognosis of young-onset *BRCA1/2* breast cancer, we assessed whether time between recent childbirth and breast cancer diagnosis is associated with increased mortality in *BRCA1/2* breast cancer patients enrolled in the Manchester UK Centre for Genomic Medicine and Family History Clinic^38^. Evaluating potential associations between recent childbirth and survival outcomes could lead to improved strategies to prevent and treat young-onset breast cancer in germline *BRCA* PV carriers.

## Methods

### Database Setting

The study population is part of a prospectively maintained database of *BRCA* pathogenic variant PV carriers at the Manchester Centre for Genomic Medicine, UK^38,39^. Women with a family or personal history of breast or ovarian cancer were referred to the Family History Clinic (FHC) and the Manchester Centre for Genomic Medicine, both founded in 1987^38^. Parity data were collected through questionnaires and detailed pedigrees administered during clinic visits.

Testing for *BRCA1/2* PVs began in 1996. Those identified with germline *BRCA1* or *BRCA2* pathogenic variants (confirmed using ACMG/AMP criteria) are the source of the study population. Patients heterozygous for pathogenic variants and their first degree relatives were entered into a dedicated database^39^. Many additional heterozygous and obligate carriers were identified by cascading^40^. Follow-up information was collected through medical record review and from the National Cancer Registration and Analysis Service. Breast cancer subtype information was obtained through abstraction of patient pathology reports.

### Ethics and Informed consent

The parent study was approved by the University of Manchester ethics review board.

Oregon Health & Science University (OHSU) received de-identified data and the research was IRB approved as exempt for the secondary-data analyses study.

### Participants

As of November 2021, a total of 1,712 unrelated families with germline *BRCA1/2* pathogenic variants and 3,588 women heterozygous for *BRCA1/2* PVs were identified in the database. After excluding n=1,654 non-cancer patients, and n=6 stage IV patients or patients with missing data on breast cancer status, a total of n=1,928 breast cancer patients with *BRCA1/2* PVs were identified (**Supplemental Figure 1**). Prophylactic mastectomy and oophorectomy surgery prior to breast cancer diagnosis has been shown to reduce breast cancer mortality among *BRCA1/2* heterozygotes^40^, and our data also show survival benefit among these individuals although not statistically significant (**Supplemental Figure 2**). Thus, we exclude n=65 patients who had oophorectomy or mastectomy before breast cancer diagnosis.

Additionally, we excluded n=50 ductal carcinoma *in situ* (DCIS); n=680 patients diagnosed at age>45 years of age; n=183 patients without diagnosis date, date of first childbirth, or date of most recent childbirth; n=40 patients diagnosed during pregnancy; and n=7 diagnosed before 1950. This resulted in a final analytical cohort of N=903 eligible non-metastatic (stage I-III) breast cancer patients with germline *BRCA1* and *BRCA2* PVs, with complete time-since-recent-childbirth data, and who were diagnosed at >15 and ≤45 years of age between 1950-2021 (**Supplemental Figure 1**). The mean (SD) follow-up time was 10.8 (9.8) years (Inter Quartile Range: 2.8-16.1 years).

Within this N=903 cohort, n=224 women were nulliparous at the time of their breast cancer diagnosis and n=10 of these cases had a 1^st^ birth after their breast cancer diagnosis. We conducted a sensitivity analysis comparing the survival difference between these 10 cases and the rest of the nulliparous cases and found no statistically significant differences (**Supplemental Figure 3**). Thus, we included these 10 cases diagnosed in nulliparous women with subsequent childbirth in the nulliparous group. Breast cancer patients were followed up to Nov 4, 2021, or until death, whichever came first. This study followed the Strengthening the Reporting of Observational Studies in Epidemiology (STROBE) reporting guidelines.

### Outcomes, Exposures, and Covariates

The primary outcome for this study is all-cause mortality. Survival duration was calculated as the time between the date of breast cancer initial diagnosis and the date of death or the date of last contact, up to the study cutoff date (Nov 4, 2021). Follow-up time was censored at 20 years.

The main exposure is the time interval between most recent childbirth and breast cancer diagnosis, using previous definitions of PPBC defined as diagnosis up to 10 years since most recent childbirth^25^. Analyses were performed on the 3-groups (nulliparous, PPBC >0-<10 years, ≥10 years since recent childbirth). Also, where sample sizes permitted, we further delineated the PPBC group into >0-<5 and 5-<10 years group as closer proximity to recent childbirth has been associated with worse prognosis in some studies^22,23^.

Covariates considered include tumor estrogen receptor (ER) status (ER-positive or ER-negative), clinical stage at breast cancer diagnosis, patient age at diagnosis, year of breast cancer diagnosis, parity, age at first full-term-birth, age at last full-term birth, *BRCA1* vs *BRCA2* PV status, and type of *BRCA* PV, e.g., copy number variants, truncating, splice site, missense PVs, and promoter alterations. *BRCA1/2* PVs were assessed with all exons sequencing before 2014 and by next generation sequencing with multiple ligation dependent probe amplification (MLPA) for whole exon deletions or duplications after 2014.

### Statistical Analyses

Chi-square tests for each categorical variable (estrogen status, tumor size, histology grade, stage, year of diagnosis, parity status, age at first full-term birth at time of diagnosis, age at last full-term birth at time of diagnosis, age at menarche, BRCA PVs, type of PVs) and the Kruskal-Wallis test for continuous variables (age at diagnosis) were conducted. The Kaplan-Meier method was used to calculate survival estimates, and the log-rank test was used to compare survival curves by time-since-recent-childbirth group. Multivariate Cox proportional hazards regression was applied to identify factors associated with the overall mortality. The proportionality assumption was tested using Schoenfeld residuals, with *BRCA* or ER status showing a non-constant hazard over time. To account for this, the data were stratified by *BRCA* PV type or ER status where appropriate. The models are based on univariate effects of the time-since-recent-childbirth interval groups followed by multivariate models that include the following covariates: age at breast cancer diagnosis, tumor stage at diagnosis, breast cancer ER status, and BRCA PV type. Diagnosis year was not included in the main adjustment because there were no significant differences in diagnosis year in association with mortality in this dataset (**Supplemental Figure 4**). Cox Proportional Hazards was used to calculate mortality hazard ratios (HRs) and 95% confidence intervals (CIs).

By definition, data for the main exposure variable, time-since-recent-childbirth, were available to all patients in the analytic cohort. However, several covariates included missing values (**Supplemental Table 1**). The distribution of data for 1) time-since-recent-childbirth (main exposure, **Supplemental Table 2a**), and 2) mortality (outcome, **Supplemental Table 2b**), were compared between patients with and without missing values for the covariates. There were no significant differences in the number of individuals with and without missing data when comparing between the time-since-recent-childbirth groups. However, patients without missing values have significant better rates of survival than patients with missing values, an observation consistent with findings from large national cancer registries^41^. Since exclusion of patients with missing variables from analyses may introduce unintended bias and underestimate breast cancer mortality^41^, patients with missing data for each covariate were considered as a distinct category.

The analyses of potential effect modifiers of the association between time-since-recent-childbirth and survival included: parity (nulliparous, 1, ≥2), age of first full-term birth (nulliparous, < 25 years old, ≥25 years old) and age at last full-term birth (nulliparous, < 25 years old, ≥25 years old). All statistical analyses were conducted using R version 4.1.1 (R Foundation) or SAS software version 9.4 (SAS Institute, Cary, NC). Tests of statistical significance were determined using two-tailed tests, and a p ≤ 0.05 was considered statistically significant.

## Results

### Patient characteristics

The analytic cohort included 903 stage I-III breast cancer cases diagnosed at age 45 years or younger (mean age=37.3 ± 5.4 years) (**Supplemental Figure 1**). The mean (SD) follow-up time was 10.8 (9.8) years (Inter Quartile Range: 2.8-16.1 years). The study participants’ demographic and clinical characteristics are summarized in **Table 1**. There were statistically significant differences in age at diagnosis, tumor histology grade, year of diagnosis, age at first full-term birth, and age at last full-term birth across the time-since-recent-childbirth interval groups. However, there were no differences in ER status, tumor size, stage, parity (between parous groups), age at menarche, distribution of *BRCA*1 or *BRCA2* PVs, or type of PV (i.e., copy number variants, truncations) across these interval groups. Further demographic and clinical characteristics of the study population, comparing *BRCA1* vs. *BRCA2* groups, are presented in **Supplemental Table 3**. Among ER-positive cases, *BRCA2* is the dominant *BRCA* PV type ranging from 62.3% among the nulliparous group to 75% among PPBC 0-<5 group; the frequency distributions of *BRCA2* among PPBC 5-<10 and parous ≥10 group are both 70.3% (P=0.56). Among ER-negative cases, *BRCA1* PVs are dominant, ranging from 74.6% in PPBC 5-<10 group to 87.3% in PPBC 0-<5 group; the frequency distribution of *BRCA1* among nulliparous group is 81.1% and among parous ≥10 group is 80% (P=0.08). These results show strong correlations between *BRCA1 and 2* PVs and ER tumor subtype, consistent with previous reports^42^. We did not observe differences in ER tumor subtypes between the time-since-recent-childbirth interval groups in *BRCA* carriers. Further, the proportions of women with *BRCA 1 vs 2* PVs did not differ between the time-since-recent-childbirth groups.

**Table 1.**
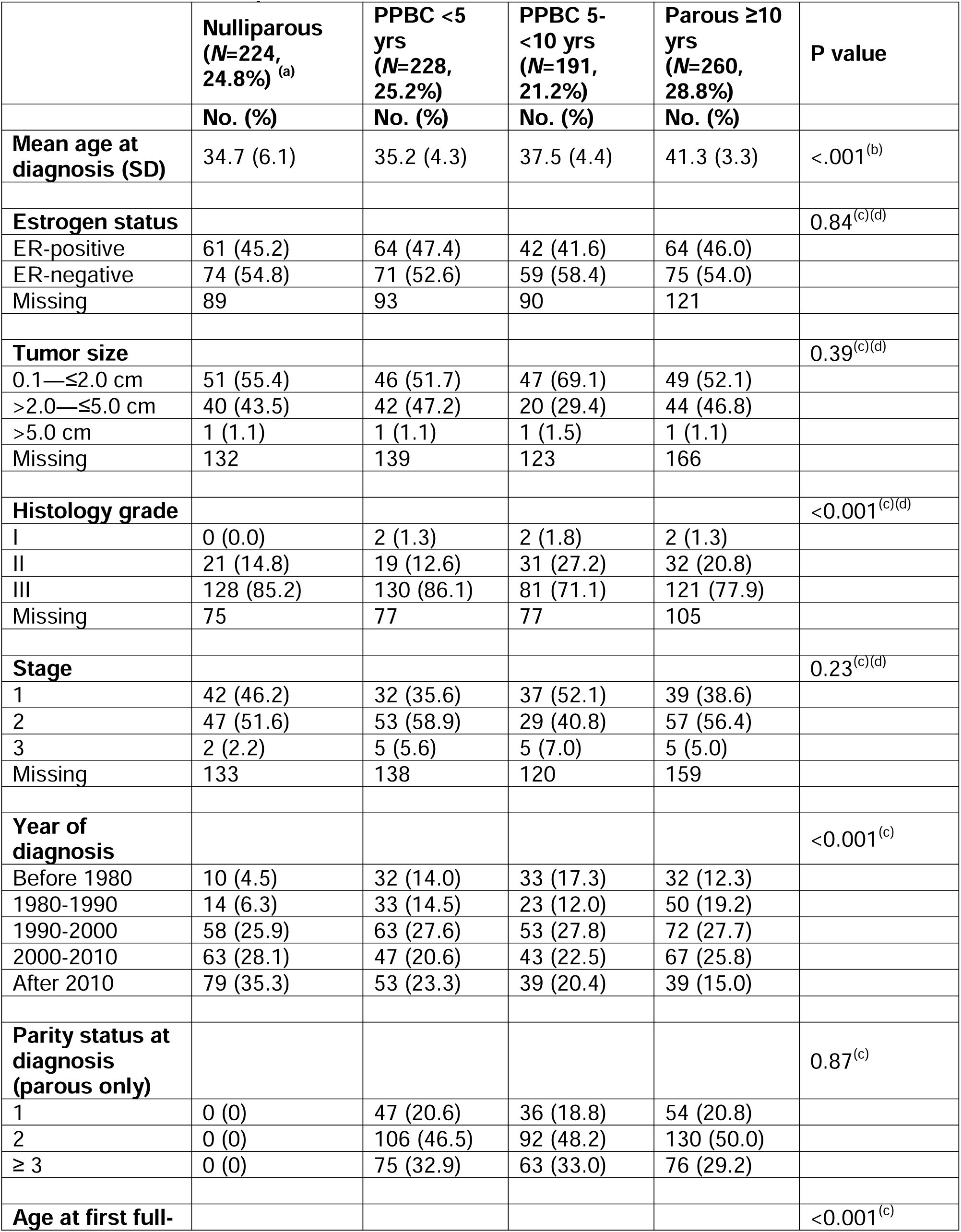
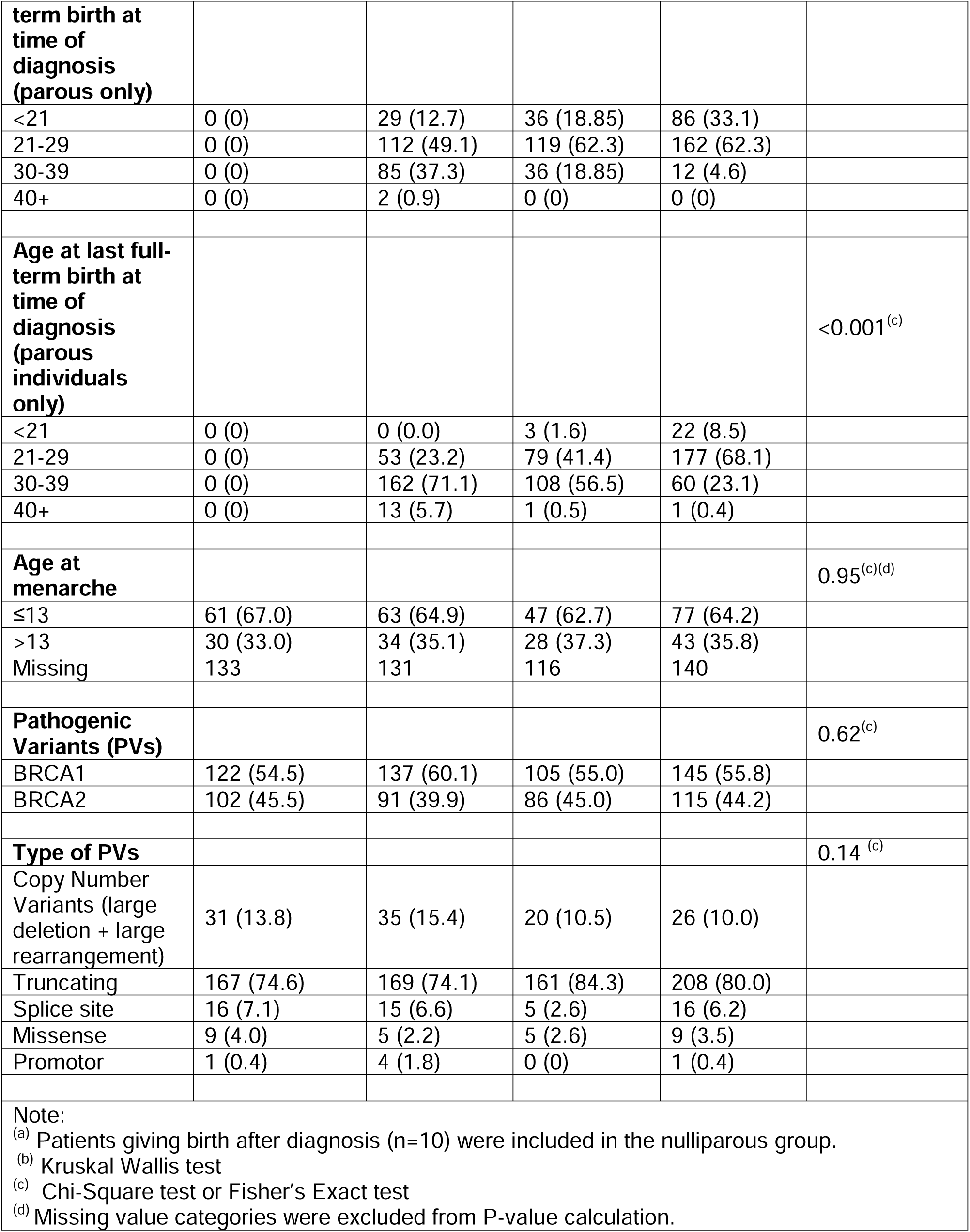
Demographic and Clinical Characteristics of the Analytic Cohort by Time-Since-Recent-Childbirth Group.

### Associations of time-since-recent-childbirth and all-cause mortality

We next evaluated the effect of time-since-recent-childbirth on overall mortality among germline *BRCA* breast cancer patients using the Kaplan-Meier method. We found a PPBC >0-<10 diagnosis (**Figure 1A**), especially PPBC 5-<10 (**Figure 1B**), had increased risk of overall mortality compared to nulliparous women. The PPBC >0-<5 group had a trend for poor prognosis compared to the nulliparous group. The parous ≥10 group had no significant difference compared to nulliparous cases (**Figure 1A & 1B**).

**Figure 1.**
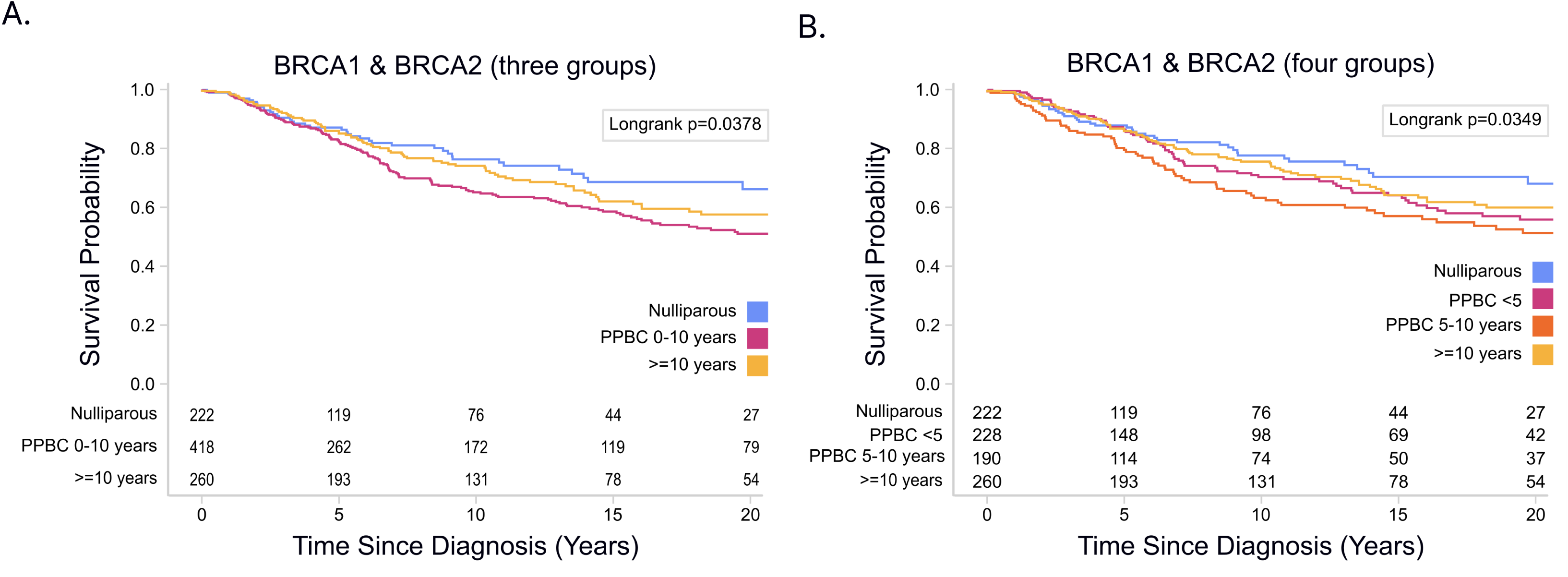
Survival outcomes by time-since-recent-childbirth for all *BRCA1* & *BRCA2* germline pathogenic variant carriers (Kaplan-Meier Curve). Different times-since-recent-childbirth groups are represented by blue (nulliparous), pink (PPBC 0-<10) [or pink (PPBC <5), dark orange (PPBC 5 - <10) for 4-group comparisons] and light orange (parous ≥10). **A.** three group comparisons of reproductive groups; **B.** four group comparisons of reproductive groups.

To determine the magnitude of the increased risk of overall mortality associated with PPBC, we conducted univariate Cox proportional hazards regression analyses. When comparing PPBC 5-<10 years vs. nulliparous, we found a 1.7 fold increased risk for overall mortality in the PPBC group (HR=1.72, CI: 1.17-2.52, p = 0.006, **Supplemental Table 4**). In multivariate analysis, this increased risk persisted after controlling for tumor stage, estrogen receptor status, PV type and age at diagnosis (HR: 1.56, CI: 1.05-2.30, p = 0.03, **Table 2**). The univariate analysis comparing PPBC 0-<5 years vs. nulliparous also showed a trend towards increased mortality [HR=1.37, CI: 0.94-2.01, p = 0.11 (**Supplemental Table 4**)], with decreased significance with adjusted analysis [HR=1.27, CI: 0.86-1.86, p = 0.23, **Table 2**)]. No difference in mortality was observed between the parous ≥ 10 years postpartum vs. nulliparous groups by univariate analysis [HR=1.23, CI: 0.85-1.79, p = 0.28 (**Supplemental Table 4**)], nor adjusted analysis [HR=1.09, CI: 0.72-1.67, p = 0.68, **Table 2**)].

**Table 2:** Multivariate Cox proportional hazard regression (HR) models for the associations between breast cancer diagnosis time since most recent childbirth and all-cause mortality.

|  | All Cases |  | ER-positive Cases |  | ER-negative Cases |  | BRCA1 Cases |  | BRCA2 Cases |  |
| --- | --- | --- | --- | --- | --- | --- | --- | --- | --- | --- |
| All Stages | HR (95% CI) | P | HR (95% CI) | P | HR (95% CI) | P | HR (95% CI) | P | HR (95% CI) | P |
| **Time since most recent childbirth (3 categories)** |  |  |  |  |  |  |  |  |  |  |
| Nulliparous<br>(n=224) | 1.00<br>(Reference) | N/A | 1.00 (Reference) | N/A | 1.00 (Reference) | N/A | 1.00 (Reference) | N/A | 1.00 (Reference) | N/A |
| PPBC 0-<10 years<br>(n=419) | 1.39 (0.98-1.97) | 0.06 | **2.28 (1.05-4.95)** | **0.04** | 1.67(0.74-3.78) | 0.22 | 1.63 (0.98-2.74) | 0.06 | 1.20 (0.75-1.95) | 0.45 |
| Parous ≥ 10 years<br>(n=260) | 1.07 (0.70-1.63) | 0.75 | 1.23 (0.41-3.67) | 0.71 | 1.45 (0.53-3.93) | 0.47 | 1.14 (0.62-2.11) | 0.67 | 1.05 (0.58-1.88) | 0.88 |
| **Time since most recent childbirth (4 categories)** |  |  |  |  |  |  |  |  |  |  |
| Nulliparous<br>(n=224) | 1.00<br>(Reference) | N/A | 1.00 (Reference) | N/A | 1.00 (Reference) | N/A | 1.00 (Reference) | N/A | 1.00 (Reference) | N/A |
| PPBC 0-<5 years<br>(n=228) | 1.27 (0.86-1.86) | 0.23 | **2.35 (1.02-5.42)** | **0.04** | 1.12 (0.44-2.83) | 0.81 | 1.39 (0.79-2.43) | 0.25 | 1.26 (0.73-2.16) | 0.41 |
| PPBC 5-<10 years<br>(n=191) | **1.56 (1.05-2.30)** | **0.03** | 2.18 (0.89-5.35) | 0.09 | **3.12 (1.22-7.97 )** | **0.02** | **2.03 (1.15-3.58)** | **0.02** | 1.15 (0.66-2.00) | 0.63 |
| Parous $\geq 10$ years<br>(n=260) | 1.09 (0.72-1.67) | 0.68 | 1.21 (0.40-3.65) | 0.73 | 1.80 (0.63-5.12) | 0.27 | 1.18 (0.64-2.18) | 0.60 | 1.03 (0.58-1.86) | 0.91 |
| Note: HR (95% CI) results are adjusted for age (continuous variable) and stage (categorical variable). |  |  |  |  |  |  |  |  |  |  |

### Associations between time-since-recent-childbirth and survival by ER status

Among ER-positive cases only, a diagnosis of PPBC >0-<10 postpartum associated with > 2 fold increased risk for overall mortality (HR=2.28, CI 1.05-4.95, p=0.04) compared to nulliparous cases (**Figure 2A**, **Table 2**). Further delineation of the postpartum cohort into PPBC 0-<5 and PPBC 5-<10 revealed that women diagnosed with ER-positive breast cancer within 0-5 years of recent childbirth had the highest increased risk for overall mortality (HR: 2.35, CI: 1.02-5.42, p = 0.04) compared to nulliparous women (**Figure 2B**, **Table 2**). Women diagnosed with ER-positive breast cancer 5-<10 years postpartum trended towards increased risk for overall mortality compared to nulliparous women (HR: 2.18, CI: 0.89-5.35, p = 0.09) (**Figure 2B**, **Table 2**). Women diagnosed with ER-positive breast cancer ≥10 years postpartum had no statistically significant difference in overall mortality compared to nulliparous women (HR: 1.21, CI: 0.40-3.65, p = 0.73) (**Figure 2B**, **Table 2**).

**Figure 2.**
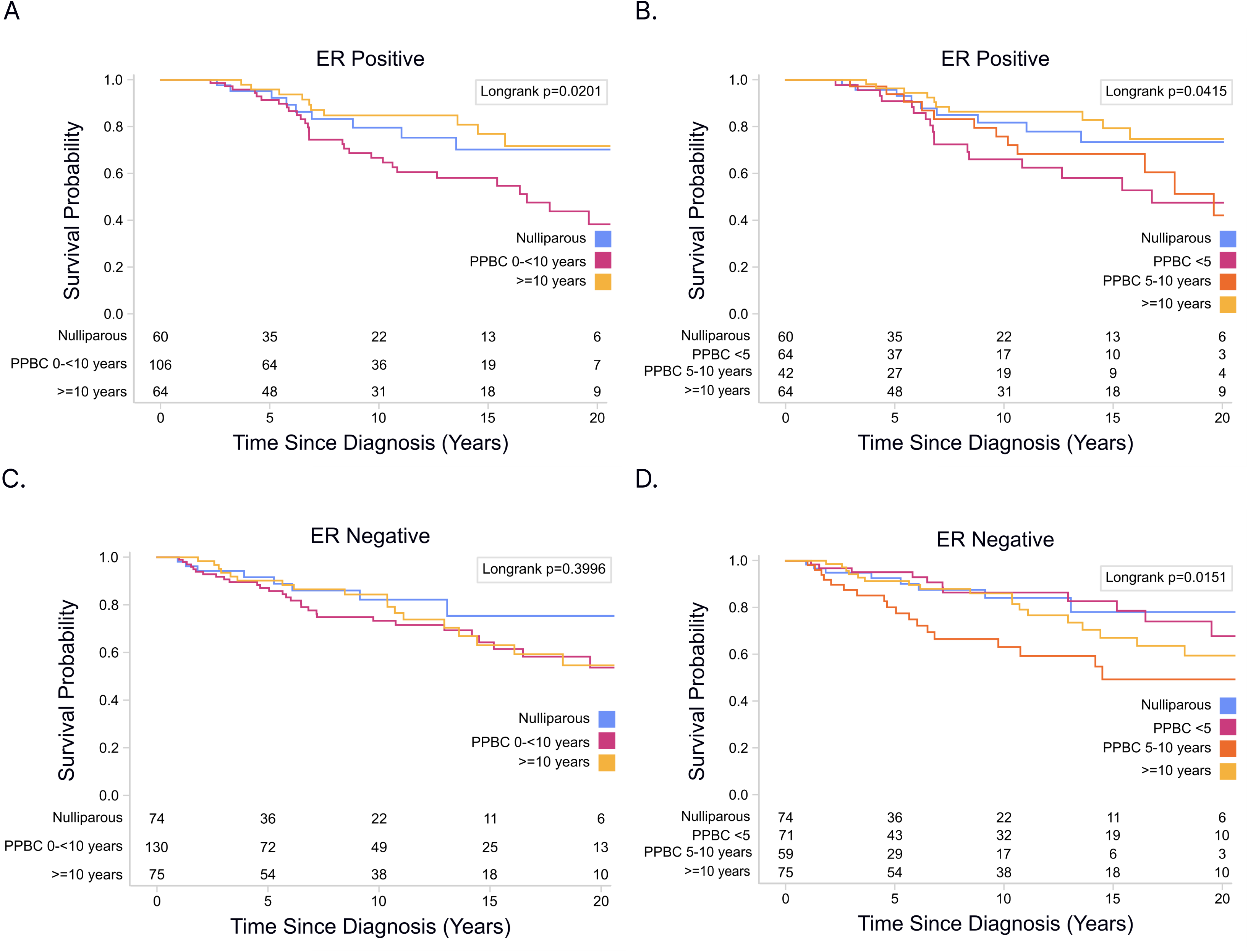
Survival outcomes by ER status and time-since-recent-childbirth (Kaplan-Meier Curve). **A.** three group comparisons of times-since-recent-childbirth groups among breast cancer patients with ER-positive tumors; **B.** four group comparisons of times-since-recent-childbirth groups among breast cancer patients with ER-positive tumors; **C.** three group comparisons of times-since-recent-childbirth groups among breast cancer patients with ER-negative tumors; **D.** four group comparisons of times-since-recent-childbirth groups among breast cancer patients with ER-negative tumors.

Among ER-negative cases, poor prognosis was not evident when PPBC was assessed as PPBC >0-<10 (HR: 1.67, CI: 0.74-3.78, p = 0.22) (**Figure 2C**, **Table 2**). However, women with ER-negative cancers diagnosed within 5-10 years of recent childbirth were three times as likely to die compared to nulliparous patients (HR: 3.12, CI: 1.22-7.97, p = 0.02) (**Figure 2D**, **Table 2**). Consistent with the postpartum risk window being transient, and not an attribute of parity per se, and similar to that observed in ER-positive disease, women diagnosed ≥10 years postpartum had no statistically significant difference for overall mortality compared to nulliparous women (HR: 1.80, CI: 0.63-5.12, p = 0.27) (**Figure 2D**, **Table 2**).

### Associations between time-since-recent-childbirth and survival by BRCA1 vs. BRCA2 status

We next sought to determine if the associations between time-since-recent-childbirth and survival differ between *BRCA1* vs *BRCA2 carriers*. In *BRCA1* cases, women diagnosed with PPBC >0-<10 trended toward overall poor prognosis compared to nulliparous women (HR: 1.63, CI: 0.98-2.74, p = 0.06, **Figure 3A**, **Table 2**). When further delineating the postpartum cohort, a 2-fold increased risk in mortality was observed in the PPBC 5-<10 group (HR: 2.03, CI: 1.15-3.58, p = 0.02, **Figure 3B**, **Table 2**). The *BRCA1* parous ≥10 years group had no statistically significant difference for overall mortality compared to nulliparous women (HR: 1.18, CI: 0.64-2.18, p = 0.27, **Figure 3B**, **Table 2**)

**Figure 3.**
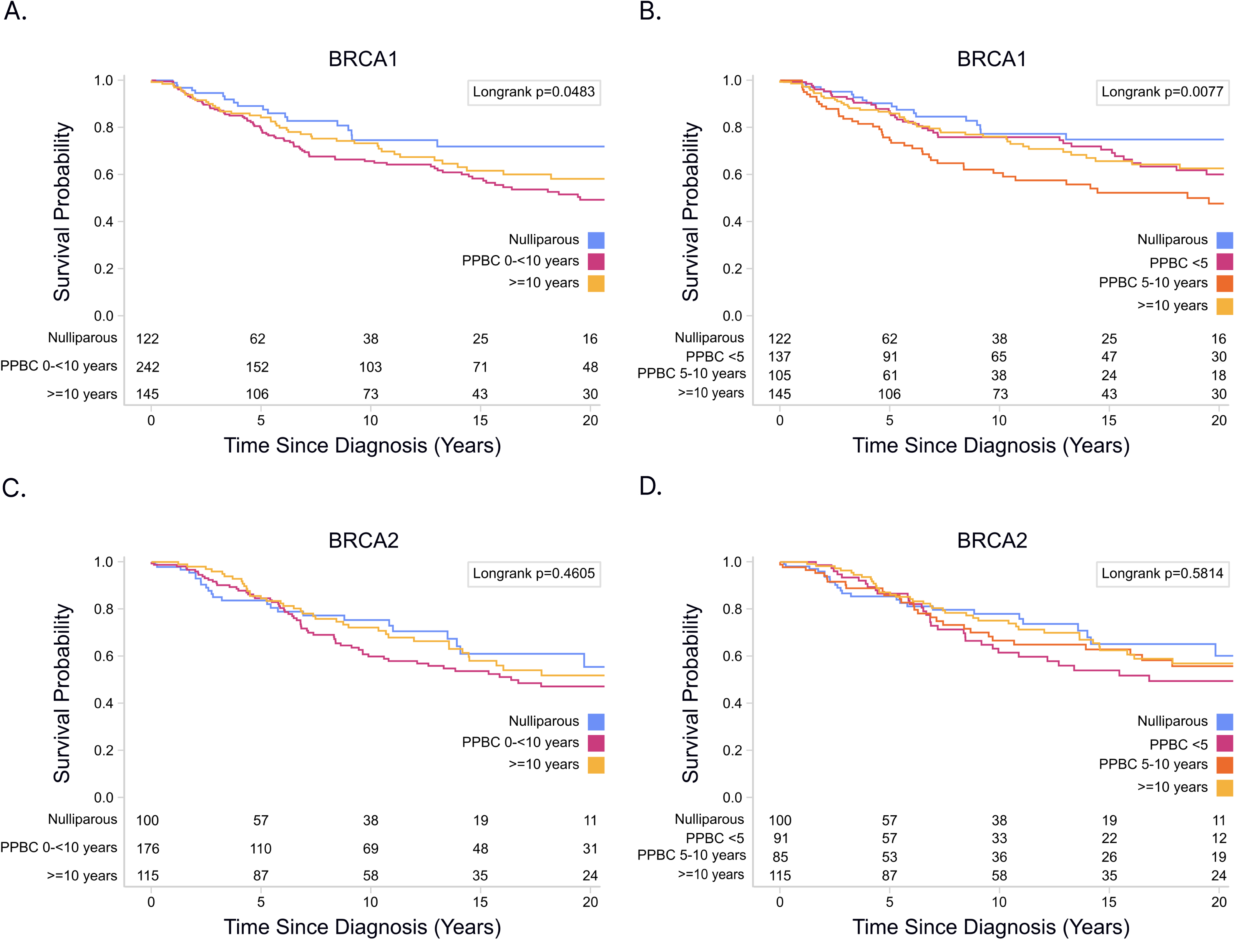
Survival outcomes by *BRCA1* vs *BRCA2* status and time-since-recent-childbirth (Kaplan-Meier Curve). **A.** three group comparisons of times-since-recent-childbirth groups among breast cancer patients with *BRCA1* mutations and; **B.** four group comparisons of times-since-recent-childbirth groups among breast cancer patients with *BRCA1* mutations and; **C.** three group comparisons of times-since-recent-childbirth groups among breast cancer patients with *BRCA2* mutations; **D.** four group comparisons of times-since-recent-childbirth groups among breast cancer patients with *BRCA2* mutations.

In *BRCA2* cases, a trend towards poorest overall survival was also observed in women diagnosed in close proximity to recent childbirth, however, in adjusted analyses, these differences did not reach significance in PPBC >0-<10 (HR: 1.20, CI: 0.75-1.95, p = 0.45, **Figure 3C**, **Table 2**), PPBC >0-<5 (HR=1.26, 95% CI, 0.73-2.16, **Figure 3D**, **Table 2**), PPBC 5-<10 (HR=1.15, 95% CI, 0.66-2.00, **Figure 3D**,**Table 2**), parous ≥10 group (HR=1.03, 95% CI, 0.58-1.86, **Figure 3D**, **Table 2**) when compared to the nulliparous group.

To further investigate how proximity to recent childbirth is a risk factor for poor prognosis in *BRCA1* carriers, but not *BRCA2* carriers, we examined overall survival differences between *BRCA1* vs. *BRCA2* cases. Survival between *BRCA1* vs. *BRCA2* cases was not significantly different overall (p=0.48), nor when stratified by time-since-recent-childbirth group (Nulliparous, p=0.21; PPBC 0-<5, p=0.14; PPBC 5-<10, p=0.22; parous ≥10, p=0.76) (**Supplemental Table 5**). One potential explanation for the poor prognosis observed in *BRCA1 PV* is that *BRCA1* is differentially regulated across a pregnancy/lactation/weaning cycle such that its loss of function during this developmental window puts the gland at increased risk for disease progression. To begin to address this question, we utilized publicly available *BRCA1/2* gene expression datasets obtained from murine models^43–45^. We found peak levels of *BRCA1* expression during the pregnancy cycle, whereas *BRCA2* was not regulated across the reproductive cycle (**Supplemental Figure 5**).

### Associations between selected reproductive variables and survival

We next evaluated if reproductive risk factors other than time-since-recent-childbirth impact breast cancer survival in *BRCA1/2* carriers. Covariates included parity (0, 1, 2, ≥3) (**Supplemental Figure 6A**), age of first full-term birth (nulliparous, <21, 21-29, ≥30) (**Supplemental Figure 6B**), and age at last full-term birth (nulliparous, <21, 21-29, ≥30) (**Supplemental Figure 6C**). None were significantly associated with overall survival: parity (p=0.15), age at first full-term birth (p=0.43), age at last full-term birth (p=0.13). In sum, these analyses identify time-since-recent-childbirth, but not the other reproductive factors, as a risk factor for reduced survival in *BRCA* carriers.

## Discussion

Women who carry a germline PV in the cancer pre-disposing genes *BRCA1 or BRCA2* have approximately 70% lifetime risk of developing breast cancer^37,46^. Numerous studies have examined reproductive risk factors that influence *BRCA1/2* breast cancer rates with the goal of reducing incidence^37,47,48^. Here we present the relationship between reproductive risk factors and survival in young women with *BRCA1/2* breast cancer with the goal of reducing mortality and increasing early detection of lethal *BRCA1/2* driven breast cancers. Among germline *BRCA1/2* PV carriers, breast cancer diagnosis within 10 years of childbirth associated with elevated all-cause mortality overall in both ER-positive and ER-negative breast cancers. Further, in this UK germline *BRCA1/2* breast cancer cohort, number of childbirths (parity), age at first full-term birth, and age at last full-term birth were not associated with increased mortality.

These data are consistent with prior studies identifying a postpartum diagnosis as an independent risk factor for breast cancer metastases, breast cancer specific death, and overall mortality, across diverse, breast cancer populations^22,23,26–30^. Further, results of this study expand the understanding of PPBC by demonstrating increased risk for mortality in germline *BRCA* PV carriers diagnosed with PPBC. This study has implications for standard of care for young-onset *BRCA1/2* breast cancer patients, as well as for genetic counseling of germline *BRCA1/2* PV carriers. Specifically, consideration for treatment escalation in ER+ PPBC, and counseling for appropriate breast cancer screening and risk reduction interventions in *BRCA1/2* carriers with recent childbirth may be warranted.

To date, the primary driver of breast cancer metastasis in PPBC has been linked to tumor extrinsic factors, i.e. the physiologically normal, but tumor supportive tissue microenvironment of the postpartum, involuting breast^49^. These pro-tumor stromal changes include the presence of activated fibroblasts^50^, pro-tumor collagen deposition^50–52^, lymphangiogenesis^53^, and immune infiltrate of immune suppressive and regulatory cells^54–58^. Evidence that these physiologic stromal changes durably alter PPBC is supported by the distinct molecular and cellular profiles observed in PPBC compared to stage and ER-subtype matched tumors diagnosed in nulliparous women^59,60^. Specifically, PPBC profiles strongly associate with normal breast involution profiles, including increased tumor collagen fibrosis, lymphovascular invasion, immune infiltrate, and gene expression profiles characterized by immunosuppression and tumor cell invasion^59,60^. Whether *BRCA* mutant PPBC tumors display a distinct molecular profile consistent with involution and disease progression, as predicted based on the poor outcomes observed in this UK cohort, remains to be determined.

We found the ER status of the tumors in this young-onset breast cancer cohort to strongly delineate between *BRCA1* and *BRCA2* carriers aligning well with previous reports of differential ER status between *BRCA1/2* carriers overall^61,62^. In our study, 77.1% of *BRCA1* PV carriers were diagnosed with ER-negative tumors, and 75.6% of *BRCA2* carriers were diagnosed with ER-positive tumors. Of note, the ratio of ER-positive to ER-negative tumors in *BRCA2* carriers mirrors the general, non-familial breast cancer population, i.e. ∼75% ER-positive and ∼25% ER-negative. One interpretation is that *BRCA2* interfaces with breast cancer downstream of factors that determine ER subtype. Conversely, *BRCA1* carriers have >3.3 fold increased probability of having ER-negative disease compared to the general breast cancer population, which suggests that loss of *BRCA1* function may be a contributing factor to the development of ER-negative breast cancer. Indeed, *BRCA1*-associated breast cancer is suggested to originate from luminal epithelial progenitors, a predominantly ER-negative cell population^63,64^. In mice, loss of *BRCA1* function inhibits the differentiation of ER-negative luminal progenitor cells into ER-positive epithelial cells^65^, and promotes the expansion of ER-negative luminal progenitors in mammary tissue^64,66,67^. Conditional knockout of *BRCA1* function results in the development of mammary tumors with characteristics similar to *BRCA1* human tumors^63^.

These results suggest that loss of *BRCA1* function may result in accumulation of ER-negative luminal progenitor cells vulnerable for oncogenic transformation, and provide a rationale for the increased incidence of ER-negative disease observed in *BRCA1* carriers.

Of note, we did not see an increase in ER-negative disease in our postpartum *BRCA1* or *BRCA2* cohorts compared to the nulliparous patients. These data suggest that increased ER-negative disease does not account for the poor prognosis of PPBC in this cohort, a result similar to that reported in other PPBC studies^22,23,28^. More research is needed to clarify potential relationships between mortality associated with close proximity to recent childbirth and breast cancer subtypes overall, and in *BRCA* carriers specifically.

Our study also finds that ER-positive and ER-negative disease have different postpartum windows of risk. Because breast cancer latency is thought to be greater than 5 years from initiation to overt cancer^68,69^, the increase in poor prognostic ER-positive cases diagnosed within 5 years of child birth is consistent with promotion of pre-existing sub-clinical tumors. The ER-negative cases had poorest prognosis if diagnosed 5-<10 years postpartum, which could indicate promotion of existing as well as initiation of breast cancer. Of note, there is evidenced for increased *BRCA1* expression during pregnancy^70,71^, consistent with a role in alveolar expansion^66,67^. During the high-proliferative window of pregnancy, loss of *BRCA1* function might exacerbate DNA damage given its critical role in DNA damage surveillance, and lead to increased oncogenic transformation. Further, receptor activator of NF-κB (RANK) and its ligand (RANK-L) play an essential role in breast development during pregnancy^72,73^ and RANK and RANK-L have been shown to promote breast cancer in *BRCA1* mutant mice^74^. These preclinical studies are consistent with the idea that *BRCA1* may have unique functions during a reproductive cycle. *BRCA1* also regulates p53-dependent gene expression^75^, and co-occurrence of somatic *TP53* PVs is more commonly observed with *BRCA1* PVs, compared to *BRCA2* PVs^76^. Further, mammary-specific deletion of TP53 and *BRCA1* leads to the development of murine mammary tumors having genomic and transcriptomic similarities to human basal-like breast cancer^77^. These potential biologic mechanisms linking *BRCA1* to lobule expansion during pregnancy may offer insights into why the peak risk window for poor prognosis among *BRCA1-*PV carriers was observed later, at 5-10 years after childbirth PPBC.

Our findings of a non-significant relationship between *BRCA2* and increased mortality risk of PPBC may suggest that *BRCA1 c*arriers are primarily responsible for the overall increase in mortality observed in the combined *BRCA1/2* PPBC cases. However, for *BRCA2* carriers, there is a trend for poorer prognosis with time-since-recent-childbirth, where survival is non-significantly poorest in the >0-<5 years since recent childbirth. Our study may be underpowered to fully investigate the impact of *BRCA2* in PPBC.

Our study also implicates a postpartum diagnosis, rather than the germline presence of BRCA1/2 PVs, as the key contributor to worse mortality in PPBC. Although *BRCA1* and *BRCA2* gene PVs represent more than 50% of all gene PVs associated with young-onset breast cancer^78^, it has been reported that young-onset breast cancer patients with *BRCA1/2* PVs have survival rates similar to young-onset breast cancer patients without these PVs^79,80^. Further, in this UK cohort, we found no significant difference in 20-year overall mortality (p=0.48) between *BRCA1* vs. *BRCA 2* PV carriers with young-onset breast cancer; while finding a diagnosis ≤10 years postpartum was associated with higher risk of mortality compared to nulliparous cases, or in women diagnosed ≥10 years after childbirth. These results suggest that proximity to recent childbirth ≤10 years before breast cancer diagnosis likely has a more pronounced impact on mortality in young-onset breast cancer than the presence of a *BRCA* germline PV.

## Strengths and Limitations

Strengths of our study include the large sample size of young-onset breast cancer patients with germline *BRCA1/2* PVs and the availability of long-term follow up data. Further, we have rigorous data on time interval between recent childbirth and breast cancer diagnosis, a variable frequently missing from many breast cancer databases including The Cancer Genome Atlas Program (TCGA) and Metabric databases. Another study strength is that we have DNA sequencing data for the *BRCA1* and *BRCA2* genes, allowing us to identify the range and nature of PVs present. Further, we have a sufficient number of combined *BRCA1 and BRCA2* PVs cases with confirmed ER status [n=231 for ER-positive; n=279 for ER-negative] to perform analysis of the mortality risk stratified by time-since-recent-childbirth group and ER subtype.

However, since we are missing ER status on 44% of our cohort, selection bias cannot be ruled out. Another study limitation is that the dataset was underpowered to conduct separate analyses for *BRCA1* vs *BRCA2* PVs carriers stratified by ER status. Further, rigorous evaluation of HER2 status by reproductive category was not possible due to the lack of HER2 clinical data. We also do not have treatment data, which could impact overall mortality if different between groups.

However, because this study comprises a geographically homogeneous population of breast cancer patients in the Northern England area, disparities in treatment approaches across groups may be minimized. Further, the wide range of diagnoses eras and available treatment may potentially affect meaningful comparisons. Another study limitation is that we do not have race/ethnicity data from the UK population, which may limit the generalization of the results to the other populations.

## Conclusions

These data suggest that germline *BRCA* PV carriers are at increased risk for all-cause mortality when breast cancer is diagnosed within 10 years of recent childbirth, compared with nulliparous women and those diagnosed greater than 10 years postpartum. These findings are similar to the general breast cancer population, where increased risk of metastasis and poor survival is observed in women diagnosed within 10 years of childbirth. For *BRCA1* patients, the risk of increased mortality is especially significant 5-10 years postpartum. This delayed risk window may suggest an interaction between *BRCA1* and a pregnancy cycle that results in initiation of new cancers, in addition to the promotion of existing, sub-clinical tumors. Further research is needed to address this possibility. In sum, consideration of the potential impact of childbirth on breast cancer outcomes in young germline *BRCA* PV carriers may improve standard of care within the realms of genetic counseling, disease prevention, and the clinic.

## Supplementary Material

Refer to Web version on PubMed Central for supplementary material

## Data Availability

All data produced in the present study are available upon reasonable request to the authors

## Acknowledgements

We thank Weston Anderson for figure design and manuscript editing.

## Author Contributions

*Concept and design*: Zhang, Schedin

*Acquisition, analysis, or interpretation of data:* Evens, Zhang, Schedin, Bernhardt,

*Drafting of the manuscript*: Zhang, Bernhardt, Schedin

*Critical revision of the manuscript for important intellectual content*: All authors

*Obtained funding*: Zhang, Schedin *Statistical analysis*: Ye, Zhang

*Supervision*: Schedin

