## Supplemental Files for "A Postpartum Breast Cancer Diagnosis Reduces Survival in Germline *BRCA* pathogenic variant Carriers"

**Supplemental Figure 1.** **Analytic cohort flowchart**

The flowchart illustrates the process to identify the final analytical cohort of N=903 eligible non-metastatic (stage I-III) breast cancer patients with germline *BRCA1* or *BRCA2* pathogenic variants (PVs). These patients had complete time-since-recent-childbirth data available during the follow-up period and were diagnosed at >15 and ≤ 45 years of age between 1950 and 2021.

All *BRCA1* and *BRCA2* pathogenic variants (PVs) carriers

(N=3,588)

Exclusion of patients without data on time between most recent childbirth and breast cancer diagnosis (n=183), diagnosed during pregnancy (n=40) and diagnosed before 1950 (n=7)

Follow-Up Average 11 years mean

Analysis

Allocation

Young-onset *BRCA1/2* breast cancer patients (stage I-III) age ≤ 45 years with study relevant parity data

(N=903)

Exclude DCIS (n=50)

Exclude age > 45 years (n=680)

Young-onset *BRCA1/2* breast cancer patients (stage I-III) age ≤ 45 years

(N=1,133)

*BRCA1/2* breast cancer patients (stage I-III) (N=1,863)

Exclusion of patients without breast cancer (n=1,654) or stage IV (n=3) or missing (n=3)

Parous cases with breast cancer

- PPBC <5 years (n=228)
- PPBC ≥5 to 10 years (n=191)
- Breast cancer ≥10 years (n=260)

Exclusion of patients with oophorectomy or mastectomy before diagnosis of cancer (n=65)

*BRCA1/2* breast cancer all patients (stage I-III) (N=1,928)

Nulliparous cases with breast cancer

(n=224)

Enrollment

Analysis

**Supplemental Figure 2:** Survival comparison between those who had vs. had not oophorectomy and mastectomy before breast cancer diagnosis (p-value =0.68)

For the 65 patients who removed oophorectomy and mastectomy before breast cancer, 6 nulliparous, 2 PPBC <5 years, 5 PPBC >=5 to 10 years, and 52 PPBC >=10.

Survival curve: removing oophorectomy and mastectomy before breast cancer vs. rest

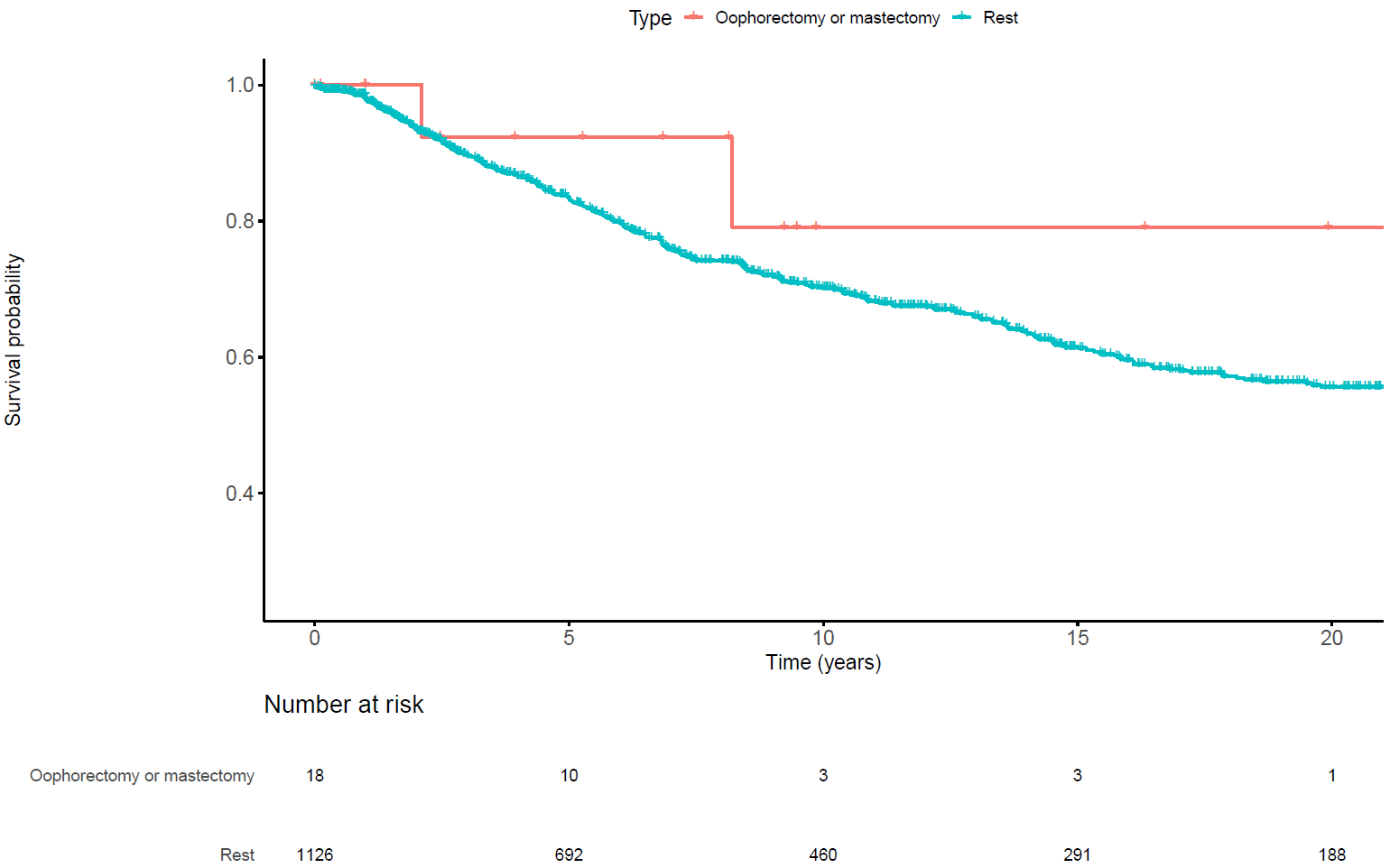

Only 18 patients in the “removing oophorectomy and mastectomy before breast cancer” group were diagnosed before 45.

**Supplemental Figure 3: Survival difference between nulliparous patients having 1^st^ childbirth after breast cancer diagnosis vs. the rest of the nulliparous patients vs. all nulliparous patients**

| **Adjustment for Multiple Comparisons for the Logrank Test** | | |
| --- | --- | --- |
| **Comparison Group** | | **Tukey_Kramer**  **P-value** |
| **1st childbirth after breast cancer diagnosis (n=10)** | **All nulliparous excluding those 10 cases (n=212)** | 0.82 |
| **1st childbirth after breast cancer diagnosis (n=10)** | **All nulliparous including those 10 cases (n=222)** | 0.95 |
| **All nulliparous excluding those 10 cases (n=212)** | **All nulliparous including those 10 cases (n=222)** | 0.98 |

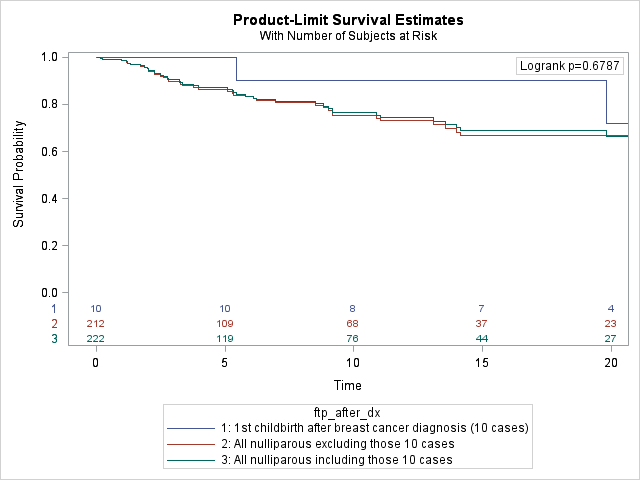

| **Reproductive Characteristics of the 10 Nulliparous Cases with Children after Diagnosis** | |
| --- | --- |
| **Parity (Parous individuals only)** | **N (%)** |
| 1 | 7 (70.0b) |
| 2 | 3 (30.0) |
| ≥ 3 | 0 (0) |
| **Age at First Full-term Birth (Parous individuals only)** |  |
| <21 | 0 (0) |
| 21-29 | 1 (10.0 ) |
| 30-39 | 8 (80.0) |
| 40+ | 1 (10.0) |
| **Age at Last Full-term Birth (Parous individuals only)** |  |
| <21 | 0 (0.0) |
| 21-29 | 1 (10.0) |
| 30-39 | 7 (70.0) |
| 40+ | 2 (20.0) |

**Supplemental Figure 4:** **Survival difference by year of diagnosis category**

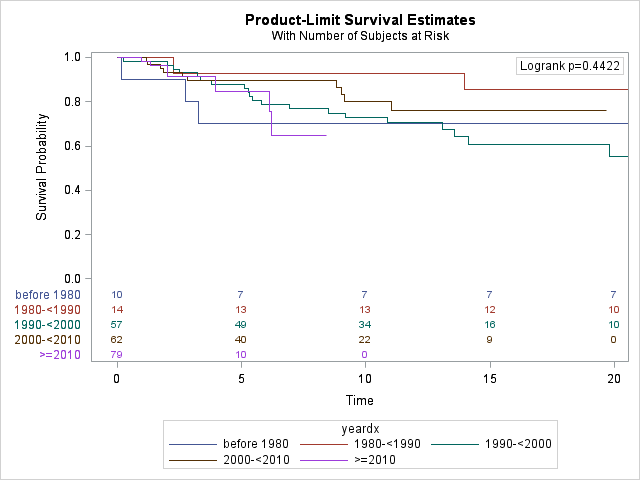

**Supplemental Figure 5: Evaluating BRCA1/2 gene expression public available data from murine mammary glands**

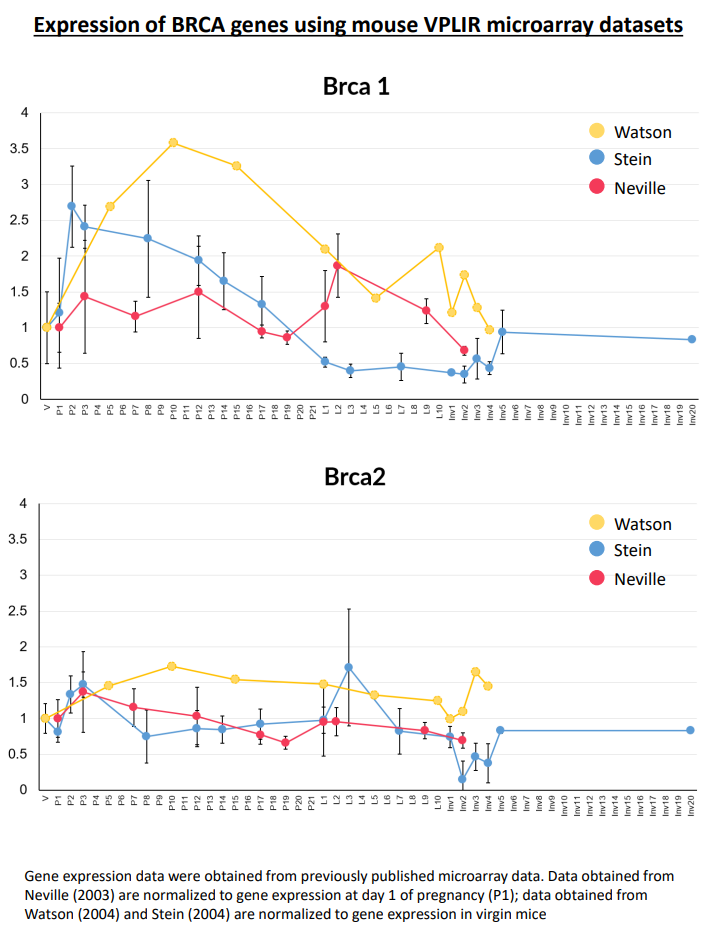

**Supplemental Figure 6. Survival outcome by reproductive variable status and time-since-recent-childbirth groups**

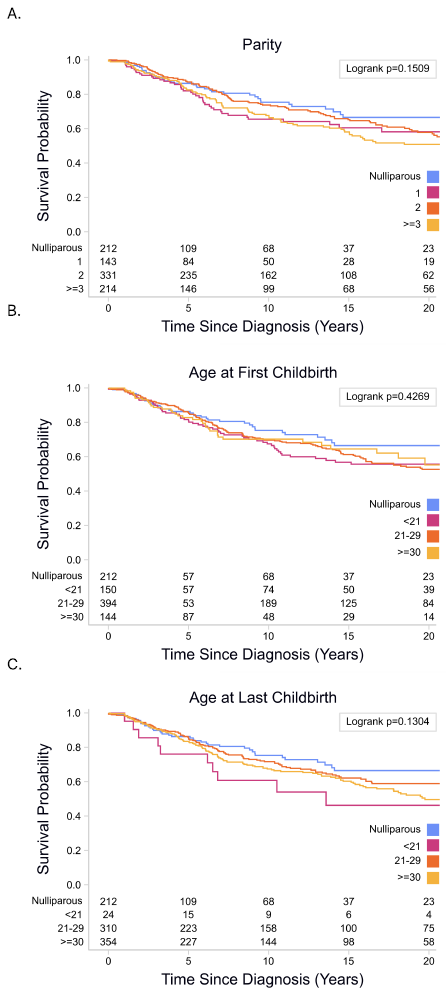

**Survival outcomes by selected reproductive variables and time-since-recent-childbirth. A. Parity:** Different parity groups are represented by blue (nulliparous), pink (parity=1), dark orange (parity=2) and light orange (parity ≥3). **B. Age at first childbirth:** Different parity groups are represented by blue (nulliparous), pink (age at first childbirth<21), dark orange (age at first childbirth 21-29) and light orange (age at first childbirth≥30). **C. Age at last childbirth:** Different parity groups are represented by blue (nulliparous), pink (age at last childbirth <21 years old), dark orange (age at last childbirth 21-29 years old) and light orange (age at last childbirth ≥30 years old)

**Supplemental Table 1. Missing Data Percentage for Variables with Missing Values**

|  | **No. (%)** |
| --- | --- |
| **Estrogen status** |  |
| ER+ | 231 (25.6) |
| ER- | 279 (30.9) |
| Missing | 393 (43.5) |
| **Tumor size** |  |
| 0.1—≤2.0 cm | 193 (21.4) |
| >2.0—≤5.0 cm | 146 (16.2) |
| >5.0 cm | 4 (0.4) |
| Missing | 560 (62.0) |
| **Histology Grade** |  |
| I | 6 (0.7) |
| II | 103 (11.4) |
| III | 460 (50.9) |
| Missing | 334 (37.0) |
| **Stage** |  |
| 1 | 150 (16.6) |
| 2 | 186 (20.6) |
| 3 | 17 (1.9) |
| Missing | 550 (60.9) |
| **Age at menarche** |  |
| ≤13 | 248 (27.5) |
| >13 | 135 (15.0) |
| Missing | 131 (57.6) |

**Supplemental Table 2a. Comparing PPBC status among individuals with missing Data vs. non-missing data by examined variables**

|  | **Nulliparous**  **(*N*=224, 24.8%) ^(a)^** | **PPBC <5**  **(*N*=228, 25.2%)** | **PPBC 5-<10 (*N*=191, 21.2%)** | **PPBC ≥10**  **(*N*=260, 28.8%)** | **P value** |
| --- | --- | --- | --- | --- | --- |
|  | **No. (%)** | **No. (%)** | **No. (%)** | **No. (%)** |  |
| **Estrogen status** |  |  |  |  | 0.26 |
| Not Missing | 135 (60.3) | 135 (59.2) | 101 (52.9) | 139 (53.5) |  |
| Missing | 89 (39.7) | 93 (40.8) | 90 (47.1) | 121 (46.5) |  |
| **Tumor size** |  |  |  |  | 0.61 |
| Not Missing | 92 (41.1) | 89 (39.0) | 68 (35.6) | 94 (36.1)) |  |
| Missing | 132 (58.9) | 139 (61.0) | 123 (64.4) | 166 (63.9) |  |
| **Histology Grade** |  |  |  |  | 0.22 |
| Not Missing | 149 (66.5) | 151 (66.2) | 114 (59.7) | 155 (59.6) |  |
| Missing | 75 (33.5) | 77 (33.8) | 77 (40.3) | 105 (40.4) |  |
| **Stage** |  |  |  |  | 0.91 |
| Not Missing | 91 (40.6) | 90 (39.5) | 71 (37.2) | 101 (38.8) |  |
| Missing | 133 (59.4) | 138 (60.5) | 120 (62.8) | 159 (61.2) |  |
| **Age at menarche** |  |  |  |  | 0.46 |
| Not Missing | 91 (40.6) | 97 (42.5) | 75 (39.3) | 120 (46.2) |  |
| Missing | 133 (59.4) | 131 (57.5) | 116 (60.7) | 140 (53.8) |  |

**Supplemental Table 2b. Comparing survival among individuals with missing Data vs. non-missing data by examined variables**

|  | **Alive** | **Death** | **P value** |
| --- | --- | --- | --- |
|  | **No. (%)** | **No. (%)** |  |
| **Estrogen status** |  |  | <0.001 |
| Not Missing | 393 (67.2) | 117 (36.8) |  |
| Missing | 192 (32.8) | 201 (63.2) |  |
| **Tumor size** |  |  | <0.001 |
| Not Missing | 261 (44.6) | 82 (25.8) |  |
| Missing | 324 (55.4) | 236 (74.2) |  |
| **Histology Grade** |  |  | <0.001 |
| Not Missing | 419 (71.6) | 150 (47.2) |  |
| Missing | 166 (28.4) | 168 (52.8) |  |
| **Stage** |  |  | <0.001 |
| Not Missing | 267 (45.6) | 86 (27.0) |  |
| Missing | 318 (59.4) | 232 (73.0) |  |
| **Age at menarche** |  |  | <0.001 |
| Not Missing | 291 (49.7) | 92 (28.9) |  |
| Missing | 294 (50.3) | 226 (71.1) |  |

**Supplemental Table 3. Demographic and Clinical Characteristics of Analytic Cohort by *BRCA* Mutation Status Group**

|  | ***BRCA1* (*N*=509, 56%) ^(a)^** | ***BRCA2* (*N*=394, 44%)** | **P value** |
| --- | --- | --- | --- |
|  | **No. (%)** | **No. (%)** |  |
| **Mean age at diagnosis (SD)** | 36.9 (5.5) | 37.8 (5.2) | 0.06^(b)^ |
| **Estrogen status** |  |  | <0.001 ^(c)(d)^ |
| ER+ | 67 (22.9) | 164 (75.6) |  |
| ER- | 226 (77.1) | 53 (24.4) |  |
| Missing | 216 | 177 |  |
| **Tumor size** |  |  | 0.84^(c)(d)^ |
| 0.1—≤2.0 cm | 102 (54.8) | 91 (58.0) |  |
| >2.0—≤5.0 cm | 82 (44.1) | 64 (40.8) |  |
| >5.0 cm | 2 (1.1) | 2 (1.3) |  |
| Missing | 323 | 237 |  |
| **Histology Grade** |  |  | <0.0001^(c)(d)^ |
| I | 1 (0.3) | 5 (2.1) |  |
| II | 18 (5.5) | 85 (35.3) |  |
| III | 309 (94.2) | 151 (62.7) |  |
| Missing | 181 | 153 |  |
| **Stage** |  |  | 0.51^(c)(d)^ |
| 1 | 87 (45.1) | 63 (39.4) |  |
| 2 | 98 (50.8) | 88 (55.0) |  |
| 3 | 8 (4.2) | 9 (5.6) |  |
| Missing | 316 | 234 |  |
| **Year of Diagnosis** |  |  | 0.77^(c)^ |
| Before 1980 | 65 (12.8) | 42 (10.7) |  |
| 1980-1990 | 66 (13.0) | 54 (13.7) |  |
| 1990-2000 | 132 (25.9) | 114 (28.9) |  |
| 2000-2010 | 125 (24.6) | 95 (24.1) |  |
| After 2010 | 121 (23.8) | 89 (22.6) |  |
| **Parity** |  |  | 0.04^(c)^) |
| 0 | 115 (22.6) | 99 (25.1) |  |
| 1 | 93 (18.3) | 51 (12.9) |  |
| 2 | 172 (33.8) | 159 (40.4) |  |
| ≥ 3 | 129 (25.3) | 85 (21.6) |  |
| **Age at first FTP** |  |  | 0.0003^(c)^ (excluding nulliparous group) |
| <21 | 93 (23.6) | 58 (19.7) |  |
| 21-29 | 224 (56.9) | 170 (57.6) |  |
| 30-39 | 74 (18.8) | 67 (22.7) |  |
| 40+ | 3 (0.8) | 0 (0.0) |  |
| **Age at last FTP** |  |  | 0.61 ^(c)^ (excluding nulliparous group) |
| <21 | 15 (3.8) | 10 (3.4) |  |
| 21-29 | 180 (45.7) | 130 (44.1) |  |
| 30-39 | 187 (47.5) | 150 (50.9) |  |
| 40+ | 12 (3.1) | 5 (1.7) |  |
| **Age at menarche** |  |  | 0.14^(c)(d)^ |
| ≤13 | 137 (61.7) | 111 (68.9) |  |
| >13 | 85 (38.3) | 50 (31.1) |  |
| Missing | 287 | 233 |  |
| **Type of mutation** |  |  | <0.001^(c)^ |
| Copy Number Variants (Large deletion + Large rearrangement) | 92 (18.1) | 20 (5.1) |  |
| Truncating | 360 (70.7) | 345 (87.5) |  |
| Splice site | 34 (6.7) | 18 (4.6) |  |
| Missense | 17 (3.3) | 11 (2.8) |  |
| Promotor | 6 (1.2) | 0 (0.0) |  |
| Note:  ^(a)^ Patients gave birth after diagnosis (n=10) were included in the nulliparous group.  ^(b)^Kruskal Wallis test  ^(c)^ Chi-Square test or Fisher’s Exact test  ^(d)^ Missing value categories were excluded from P-value calculation. | | | |

**Supplemental Table 4. Unadjusted and age-adjusted Cox proportional hazard regression (HR) models for the associations between breast cancer diagnosis time since First childbirth status and survival**

|  | **Unadjusted** | | | **Age-adjusted** | | |
| --- | --- | --- | --- | --- | --- | --- |
| Time since First childbirth | HR (95% CI) | P | Overall P | HR (95% CI) | P | Overall P |
| **All Stages** |  |  |  |  |  |  |
| Nulliparous | 1.00 (Reference) | N/A | **0.048** | 1.00 (Reference) | N/A | **0.037** |
| PPBC 0-<5 years | 1.37 (0.94-2.01) | 0.11 |  | 1.36 (0.93-1.99) | 0.12 |  |
| Parous 5-<10 years | 1.72 (1.17-2.52) | **0.006** |  | 1.64 (1.11-2.42) | **0.01** |  |
| Parous ≥ 10 years | 1.23 (0.85-1.79) | 0.28 |  | 1.09 (0.72-1.66) | 0.69 |  |
| **Stages I & II** |  |  |  |  |  |  |
| Nulliparous | 1.00 (Reference) | N/A | **0.025** | 1.00 (Reference) | N/A | **0.025** |
| PPBC 0-<5 years | 1.09 (0.54-2.18) | 0.81 |  | 1.11 (0.55-2.24) | 0.76 |  |
| Parous 5-<10 years | 1.90 (0.98-3.68) | 0.058 |  | 2.04 (1.02-4.04) | **0.04** |  |
| Parous ≥ 10 years | 0.70 (0.35-1.40) | 0.31 |  | 0.83 (0.37-1.90) | 0.66 |  |
| **BRCA1** |  |  |  |  |  |  |
| Nulliparous | 1.00 (Reference) | N/A | **0.011** | 1.00 (Reference) | N/A | **0.01** |
| PPBC 0-<5 years | 1.48 (0.84-2.58) | 0.17 |  | 1.42 (0.81-2.49) | 0.17 |  |
| Parous 5-<10 years | 2.41 (1.39-4.19) | **0.002** |  | 2.22 (1.26-3.89) | **0.006** |  |
| Parous ≥ 10 years | 1.48 (0.85-2.56) | 0.17 |  | 1.20 (0.65-2.22) | 0.57 |  |
| **BRCA2** |  |  |  |  |  |  |
| Nulliparous | 1.00 (Reference) | N/A | 0.613 | 1.00 (Reference) | N/A | 0.59 |
| PPBC 0-<5 years | 1.38 (0.82-2.35) | 0.23 |  | 1.39 (0.82-2.35) | 0.23 |  |
| Parous 5-<10 years | 1.19 (0.69-2.05) | 0.54 |  | 1.18 (0.68-2.05) | 0.56 |  |
| Parous ≥ 10 years | 1.04 (0.63-1.74) | 0.87 |  | 1.02 (0.57-1.83) | 0.94 |  |
| **ER+** |  |  |  |  |  |  |
| Nulliparous | 1.00 (Reference) | N/A | **0.046** | 1.00 (Reference) | N/A | **0.046** |
| PPBC 0-<5 years | 2.15 (0.96-4.84) | 0.06 |  | 2.19 (0.97-4.92) | 0.06 |  |
| Parous 5-<10 years | 1.77 (0.74-4.19) | 0.20 |  | 1.90 (0.78-4.62) | 0.16 |  |
| Parous ≥ 10years | 0.81 (0.33-1.99) | 0.64 |  | 1.00 (0.35-2.90) | 1.00 |  |
| **ER-** |  |  |  |  |  |  |
| Nulliparous | 1.00 (Reference) | N/A | **0.027** | 1.00 (Reference) | N/A | **0.027** |
| PPBC 0-<5 years | 1.03 (0.42-2.57) | 0.95 |  | 1.07 (0.43-2.70) | 0.88 |  |
| Parous 5-<10 years | 2.83 (1.23-6.52) | **0.014** |  | 3.13 (1.26-7.74) | **0.014** |  |
| Parous ≥ 10 years | 1.48 (0.64-3.40) | 0.36 |  | 1.74 (0.63-4.77) | 0.28 |  |

**Supplemental Table5: Log-Rank Test Comparing BRCA status (BRCA1 vs. BRCA2) and all-cause mortality**

|  | P |
| --- | --- |
| All | 0.48 |
| Nulliparous | 0.21 |
| PPBC 0-<5 years | 0.14 |
| PPBC 5-<10 years | 0.22 |
| Parous ≥ 10 years | 0.76 |

**Supplemental Table. STROBE Table**

| **Item** | **Item** | **STROBE recommendations** | **Reported on page #** |
| --- | --- | --- | --- |
| **Title and**  **abstract** | 1 | (a) Indicate the study’s design with a commonly used term in the title or the abstract.  (b) Provide in the abstract an informative and balanced summary of what was done and what was found. | **1 and 4** |
| **Introduction** |  |  |  |
| Background rationale | 2 | Explain the scientific background and rationale for the investigation being reported. | **7** |
| Objectives | 3 | State specific objectives, including any pre-specified hypotheses. | **8** |
| **Methods** |  |  |  |
| Study design | 4 | Present key elements of study design early in the paper. | **8** |
| Settings | 5 | Describe the setting, locations, and relevant dates, including periods of recruitment, exposure, follow-up, and data collection. | **8** |
| Participants | 6 | a) Cohort study—Give the eligibility criteria, and the sources and methods of selection of participants. Describe methods of follow-up.  Case-control study—Give the eligibility criteria, and the sources and methods of case ascertainment and control selection. Give the rationale for the choice of cases and controls.  Cross-sectional study—Give the eligibility criteria, and the sources and methods of selection of participants.  (b) Cohort study—For matched studies, give matching criteria and number of exposed and unexposed.  Case-control study—For matched studies, give matching criteria and the number of controls per case. | **9** |
| Variables | 7 | Clearly define all outcomes, exposures, predictors, potential confounders, and effect modifiers. Give diagnostic criteria, if applicable. | **10** |
| Data sources - measurements | 8 | For each variable of interest, give sources of data and details of methods of assessment (measurement).Describe comparability of assessment methods if there is more than one group. | **8-10** |
| Bias | 9 | Describe any efforts to address potential sources of bias. | **N/A** |
| Study Size | 10 | Explain how the study size was arrived at. | **9** |
| Quantitative variables | 11 | Explain how quantitative variables were handled in the analyses. If applicable, describe which groupings were chosen and why. | **10-11** |
| Statistical  Methods | 12 | (a) Describe all statistical methods, including those used to control for confounding  (b) Describe any methods used to examine subgroups and interactions.  (c) Explain how missing data were addressed.  (d) Cohort study—If applicable, explain how loss to follow-up was addressed.  Case-control study—If applicable, explain how matching of cases and controls was addressed.  Cross-sectional study—If applicable, describe analytical methods taking account of sampling strategy.  (e) Describe any sensitivity analyses. | **10-12** |
| **Results** |  |  |  |
| Participants | 13 | (a) Report the numbers of individuals at each stage of the study—e.g., numbers potentially eligible, examined for eligibility, confirmed eligible, included in the study, completing follow-up, and analyzed.  (b) Give reasons for non-participation at each stage.  (c) Consider use of a flow diagram. | **10** |
| Descriptive data | 14 | (a) Give characteristics of study participants (e.g., demographic, clinical, social) and information on exposures and potential confounders  (b) Indicate the number of participants with missing data for each variable of interest  (c) Cohort study—Summarize follow-up time (e.g., average and total amount) | **9&12** |
| Outcome data | 15 | Cohort study—Report numbers of outcome events or summary measures over time.  Case-control study—Report numbers in each exposure category, or summary measures of exposure.  Cross-sectional study—Report numbers of outcome events or summary measures. | **10&12** |
| Main results | 16 | (a) Give unadjusted estimates and, if applicable, confounder-adjusted estimates and their precision (e.g., 95% confidence interval).  Make clear which confounders were adjusted for and why they were included.  (b) Report category boundaries when continuous variables were categorized.  (c) If relevant, consider translating estimates of relative risk into absolute risk for a meaningful time period. | **10-15** |
| Other analyses | 17 | Report other analyses done—e.g., analyses of subgroups and interactions and sensitivity analyses. |  |
| **Discussion** |  |  |  |
| Key results | 18 | Summarize key results with reference to study objectives. | **16** |
| Limitation | 19 | Discuss limitations of the study, taking into account sources of potential bias or imprecision. Discuss both direction and magnitude of any potential bias. | **20** |
| Interpretation | 20 | Give a cautious overall interpretation of results considering objectives, limitations, multiplicity of analyses, results from similar studies, and other relevant evidence. | **16-20** |
| Generalizability | 21 | Discuss the generalizability (external validity) of the study results. | **20** |
| **Other information** |  |  |  |
| Funding | 22 | Give the source of funding and the role of the funders for the present study and, if applicable, for the original study on which the present article is based. | **21** |
| *Ethics* |  |  |  |
| *Supplementary material* |  |  |  |
